# Breakthrough SARS-CoV-2 Infections after Vaccination in North Carolina

**DOI:** 10.1101/2021.10.10.21264812

**Authors:** Diane Uschner, Matthew Bott, Michele Santacatterina, Mihili Gunaratne, Lida M. Fette, Brian Burke, Greg Strylewicz, Sharon L. Edelstein, William H Lagarde, Kristen Miller, William S. Weintraub, Joseph Keating, John Schieffelin, Joshua Yukich, Hazel Tapp, Amina Ahmed, Andrea A. Berry, Iqra Munawar, Austin Lyles Seals, John Williamson, David Herrington, John W. Sanders, Michael Runyon, for the COVID-19 Community Research Partnership

## Abstract

**Importance:** Real-world data are needed to assess incidence and factors associated with breakthrough SARS-CoV-2 infections following vaccination.

**Objective:** Estimate incidence of breakthrough infections and assess associations with risk factors using self-reported data from a large NC population sample.

**Design:** Prospective observational cohort study utilizing daily online survey data to capture information about COVID-19 symptoms, testing, and vaccination status.

**Setting:** Six health care systems in North Carolina with data collected between January 15, 2021 and September 24, 2021.

**Participants:** Adult study participants who reported full vaccination with a COVID-19 mRNA or J&J non-replicating viral vector vaccine (n =16,020).

**Exposures:** Potential community exposure to SARS-CoV-2.

**Main Outcome and Measures:** Self-reported breakthrough infection.

**Results:** SARS-CoV-2 infection after vaccination was self-reported in 1.9% of participants, with an incidence rate of 7.3 per 100,000 person-years. Younger age (45-64 vs. 18-44: HR (95% CI) = 0.65 (0.51 - 0.82); 65+ vs. 18-44: HR (95% CI) = 0.59 (0.39 - 0.90)), and vaccination with J&J Ad26.COV2.S were associated with a higher risk of breakthrough infection compared to vaccination with Pfizer BNT162b2 (Ad26.COV2.S vs. BNT162b2: HR (95% CI) = 2.23 (1.40 - 3.56)), while participants vaccinated with mRNA-1273 (mRNA-1273 vs. BNT162b2: HR (95% CI) = 0.69 (0.50 – 0.96) and those residing in urban counties experienced a lower rate of SARS-CoV-2 breakthrough infection compared with those from suburban (HR (95% CI) = 1.39 (1.01 – 1.90) or rural (HR (95% CI) = 1.57 (1.16 – 2.11) counties. There was no significant association between breakthrough infection and participant sex, race, healthcare worker status, prior COVID-19 infection, routine mask use, or overall vaccination rate in the county of residence.

**Conclusions and Relevance:** This NC community-based observational study showed that the proportion of the cohort who self-report breakthrough SARS-CoV-2 infections was 7.3 events per 100,000 person-years. Younger adults, those vaccinated with J&J Ad26.COV2.S, and those residing in suburban or rural counties were at higher risk of breakthrough infections and should be targeted for additional risk mitigation strategies to decrease community transmission.

**Trial Registration:** The COVID-19 Community Research Partnership is listed in clinicaltrials.gov (NCT04342884).

**Key Points:** *Question:* What are the characteristics of those with breakthrough infections after SARS-CoV-2 vaccination in North Carolinaã

*Findings:* In this NC-based observational study of 16,020 participants, 1.9% self-reported a positive SARS-CoV-2 viral test at least 2 weeks following full vaccination, reflecting an event rate of 7.3 infections per 100,000 person years. Rates were higher among younger participants, participants from more rural areas in North Carolina, and those vaccinated with J&J Ad26.COV2.S.

*Meaning:* Our results show a relatively low rate of COVID-19 infection following full vaccination. Younger adults and those vaccinated with J&J Ad26.COV2.S should be targeted for additional risk mitigation strategies.

## Introduction

Despite the remarkable clinical trial efficacy^1-3^ and real-world effectiveness of the SARS-CoV-2 vaccines,^4^ breakthrough infections do occur, potentially with significant clinical and public health consequences.^5-7^ While vaccine effectiveness appears to wane over time,^8,9^ studies of breakthrough infection have focused largely on subsequent healthcare utilization and clinical outcomes.^10,11^ Little is known about the risk factors associated with breakthrough infection such as age, race, sex, community exposure and behavior, and history of prior COVID-19 infection. A better understanding of these factors may allow identification of a subset of vaccinated persons at higher risk of breakthrough infection for whom additional risk mitigation strategies may be needed. This investigation characterizes the overall incidence and risk factors associated with breakthrough infection among fully vaccinated participants in the North Carolina (NC) COVID-19 Community Research Partnership (CRP).

## Methods

The NC CRP is a prospective, observational cohort study that solicited participants for COVID-19 symptoms, test results, vaccination status and risk behavior via daily email or text surveys. Adults 18 years and older were enrolled between April 13, 2020 and August 14, 2021 at six NC healthcare systems (http://www.covid19communitystudy.org/). This study was approved by the Wake Forest School of Medicine Institutional Review Board. Participants consented to allow access to their electronic health records (EHR). Demographic data and healthcare worker status were collected at enrollment. Counties of residence were classified as urban, suburban or rural based on population density estimates.^12^ Symptoms of COVID-19 included fever, chills, cough, shortness of breath, fatigue, muscle pain, headache, loss of taste/smell, sore throat, congestion/runny nose, nausea/vomiting, and diarrhea. We defined SARS-CoV-2 vaccination as participant self-report of receiving two doses of either the Pfizer BioNTech BNT162b2 or Moderna mRNA-1273 vaccine, or one dose of the Janssen (Johnson & Johnson, J&J) non-replicating viral vector vaccine Ad26.COV2.S. Vaccination status was confirmed for the subset of participants with accessible EHR data. The primary outcome was weeks until first self-reported infection (positive SARS-CoV-2 antigen or nucleic acid amplification test) occurring ≥ 14 days after vaccination (between January 15, 2021 and September 24, 2021). The secondary outcome was time to symptomatic infection defined as one or more self-reported symptom suggestive of COVID-19 ± 3 days from the date of a positive test. General mask usage as a proxy for COVID-19 risk mitigation behavior was defined as wearing a face mask 90% or more of the time when meeting people outside the household in the 14 days after vaccination.

## Statistical methods

Descriptive statistics are presented as absolute and relative frequencies for categorical variables and median and inter-quartile range (IQR) for follow-up time. Cumulative incidence was estimated as 1 minus the Kaplan-Meier curve. Unadjusted cumulative event rates were estimated as number of events by person time. Cox proportional hazards models were used to estimate unadjusted and adjusted hazard ratios with 95% confidence intervals (CI). Only main effects were considered in the analysis. Three participants who did not specify their sex at enrollment were excluded from analysis. There were no other missing data. All variables of interest were selected a-priori and were used as covariates in the multivariable model. Exploratory data analysis highlighted the large effect of vaccination quarter, likely related to the non-uniform effect of calendar time on risk for breakthrough due to the delta SARS-CoV-2 variant (eFigure 1). Accordingly, univariate and multivariate analyses were adjusted for vaccination quarter before estimating HRs for breakthrough infection after vaccination. Analyses were performed using R (V.4.0.3, R Foundation for Statistical Computing).

## Results

Of 16,020 eligible participants, 310 (1.9%) reported a positive SARS-CoV-2 test after vaccination (eFigure 2). The event rate was 7.3 breakthrough infections per 100,000 person-years. Infections were symptomatic in 286 (92%) cases. Population characteristics are summarized in Table 1. Median and interquartile follow-up times following full vaccination were 24.0, IQR = (17–28.4) weeks among infected participants and 23.6, IQR = (17.4-29.9) weeks among uninfected participants. The cumulative incidence of breakthrough infection was 5.2% at 34 weeks following full vaccination (Figure 1). In the multivariable analysis (Table 1), age 45 and older was associated with lower risk of breakthrough infection; HR (95% CI) of age 45-64 vs. 18-44 was 0.65 (0.51 - 0.82); age 65+ vs. 18-44 was 0.59 (0.39 - 0.90). Compared to those vaccinated with BNT162b2, participants vaccinated with Ad26.COV2.S had a higher risk of breakthrough infection; HR (95% CI) = 2.23 (1.40 - 3.56) and those vaccinated with mRNA-1273 had a lower risk of breakthrough infection; HR (95% CI) = 0.69 (0.50 – 0.96). Participants from rural and suburban counties had a higher risk of breakthrough infections; HR (95% CI) of suburban vs. urban was 1.39 (1.01 – 1.90); rural vs. urban was 1.57 (1.16 - 2.11). There was no association of breakthrough infection with sex, race/ethnicity, healthcare worker status, prior self-reported COVID-19 infection, general mask usage, or vaccination rate in the county of residence.

**Table 1.** SARS-CoV-2 Infection rate after vaccination per 100,000 person-years overall and by risk factors.

| Characteristics | N (%) | # Of Events (%) | Event Rate (95% CI) | Univariate<sup>a</sup> |  | Multivariate<sup>b</sup> |  |
| --- | --- | --- | --- | --- | --- | --- | --- |
|  |  |  |  | Hazard Ratio (95% CI) | P-value | Hazard Ratio (95% CI) | P-value |
| **Age** |  |  |  |  |  |  |  |
| 18-44 (*Reference group*) | 5250 (33%) | 133 (2.5%) | 10.3 (8.7 - 12.2) |  |  |  |  |
| 45-64 | 7585 (47%) | 135 (1.8%) | 6.7 (5.7 - 8) | 0.64 (0.5 - 0.81) | **0.0003** | 0.65 (0.51 - 0.82) | **0.0004** |
| 65+ | 3185 (20%) | 42 (1.3%) | 4.4 (3.3 - 6) | 0.49 (0.34 - 0.7) | **0.0001** | 0.59 (0.39 - 0.89) | **0.0129** |
| **Sex** |  |  |  |  |  |  |  |
| Female (*Reference group*) | 11794 (74%) | 230 (2%) | 7.3 (6.4 - 8.3) |  |  |  |  |
| Male | 4226 (26%) | 80 (1.9%) | 7.2 (5.8 - 8.9) | 1.01 (0.78 - 1.3) | 0.9452 | 1.14 (0.88 - 1.48) | 0.3130 |
| **Race/Ethnicity** |  |  |  |  |  |  |  |
| Non-Hispanic White (*Reference group*) | 15054 (94%) | 294 (2%) | 7.3 (6.5 - 8.2) |  |  |  |  |
| Non-Hispanic Black | 401 (3%) | 3 (0.7%) | 3.2 (1 - 9.9) | 0.5 (0.16 - 1.55) | 0.2300 | 0.44 (0.14 - 1.38) | 0.1613 |
| Hispanic | 172 (1%) | 5 (2.9%) | 12.9 (5.4 - 31) | 1.85 (0.76 - 4.47) | 0.1746 | 1.52 (0.62 - 3.69) | 0.3580 |
| Non-Hispanic Other | 393 (2%) | 8 (2%) | 8.5 (4.2 - 16.9) | 1.2 (0.59 - 2.42) | 0.6100 | 1.03 (0.51 - 2.1) | 0.9307 |
| **Healthcare Worker Status** |  |  |  |  |  |  |  |
| No (*Reference group*) | 10559 (66%) | 158 (1.5%) | 5.9 (5.1 - 6.9) |  |  |  |  |
| Yes | 5461 (34%) | 152 (2.8%) | 9.6 (8.2 - 11.3) | 1.53 (1.16 - 2.02) | **0.0029** | 1.33 (0.95 - 1.85) | 0.0965 |
| **Vaccination Brand** |  |  |  |  |  |  |  |
| Pfizer BNT162b2 (*Reference group*) | 11199 (70%) | 242 (2.2%) | 7.9 (7 - 9) |  |  |  |  |
| Moderna mRNA-1273 | 3997 (25%) | 47 (1.2%) | 4.6 (3.5 - 6.2) | 0.68 (0.5 - 0.94) | **0.0187** | 0.69 (0.5 - 0.96) | **0.0281** |
| Janssen (Johnson & Johnson) Ad26.COV2.S | 824 (5%) | 21 (2.5%) | 11.4 (7.5 - 17.5) | 2.13 (1.34 - 3.37) | **0.0013** | 2.23 (1.4 - 3.56) | **0.0007** |
| **Prior COVID Infection** |  |  |  |  |  |  |  |
| No (*Reference group*) | 15359 (96%) | 298 (1.9%) | 7.3 (6.5 - 8.2) |  |  |  |  |
| Yes | 661 (4%) | 12 (1.8%) | 7.4 (4.2 - 13) | 1.03 (0.58 - 1.83) | 0.9323 | 0.94 (0.53 - 1.68) | 0.8382 |
| **Vaccination Rate in County of Residence** |  |  |  |  |  |  |  |
| < 60% (*Reference group*) | 13617 (85%) | 258 (1.9%) | 7.1 (6.3 - 8) |  |  |  |  |
| ≥ 60% | 2403 (15%) | 52 (2.2%) | 8.3 (6.3 - 10.9) | 1.2 (0.89 - 1.61) | 0.2404 | 1.31 (0.95 - 1.79) | 0.0966 |
| **County Classification** |  |  |  |  |  |  |  |
| Urban (*Reference group*) | 10729 (67%) | 195 (1.8%) | 6.8 (5.9 - 7.8) |  |  |  |  |
| Suburban | 2366 (15%) | 52 (2.2%) | 8.5 (6.4 - 11.1) | 1.25 (0.92 - 1.7) | 0.1539 | 1.39 (1.01 - 1.9) | **0.0407** |
| Rural | 2925 (18%) | 63 (2.2%) | 8.2 (6.4 - 10.6) | 1.27 (0.96 - 1.69) | 0.0942 | 1.57 (1.16 - 2.11) | **0.0033** |
| **Mask Usage<sup>c</sup>** |  |  |  |  |  |  |  |
| ≤ 90% (*Reference group*) | 10186 (64%) | 198 (1.9%) | 7.4 (6.4 - 8.5) |  |  |  |  |
| > 90% | 5834 (36%) | 112 (1.9%) | 7.1 (5.9 - 8.6) | 0.91 (0.72 - 1.15) | 0.4165 | 0.87 (0.68 - 1.1) | 0.2348 |
| **Total Number of Participants** | 16020 (100%) | 310 (1.9%) | 7.3 (6.5 - 8.1) |  |  |  |  |

<sup>a</sup> Hazard ratios for individual variables after adjusting for vaccine quarter (e.g. Jan-Mar, Apr-Jun, Jul-Sept)<sup>b</sup> Includes all variables in this table and is adjusted for vaccine quarter (e.g. Jan-Mar, Apr-Jun, Jul-Sept)<sup>c</sup> Mask usage in the 2 weeks following vaccination as a proxy for general COVID-19 risk mitigation behavior

**Figure 1.**
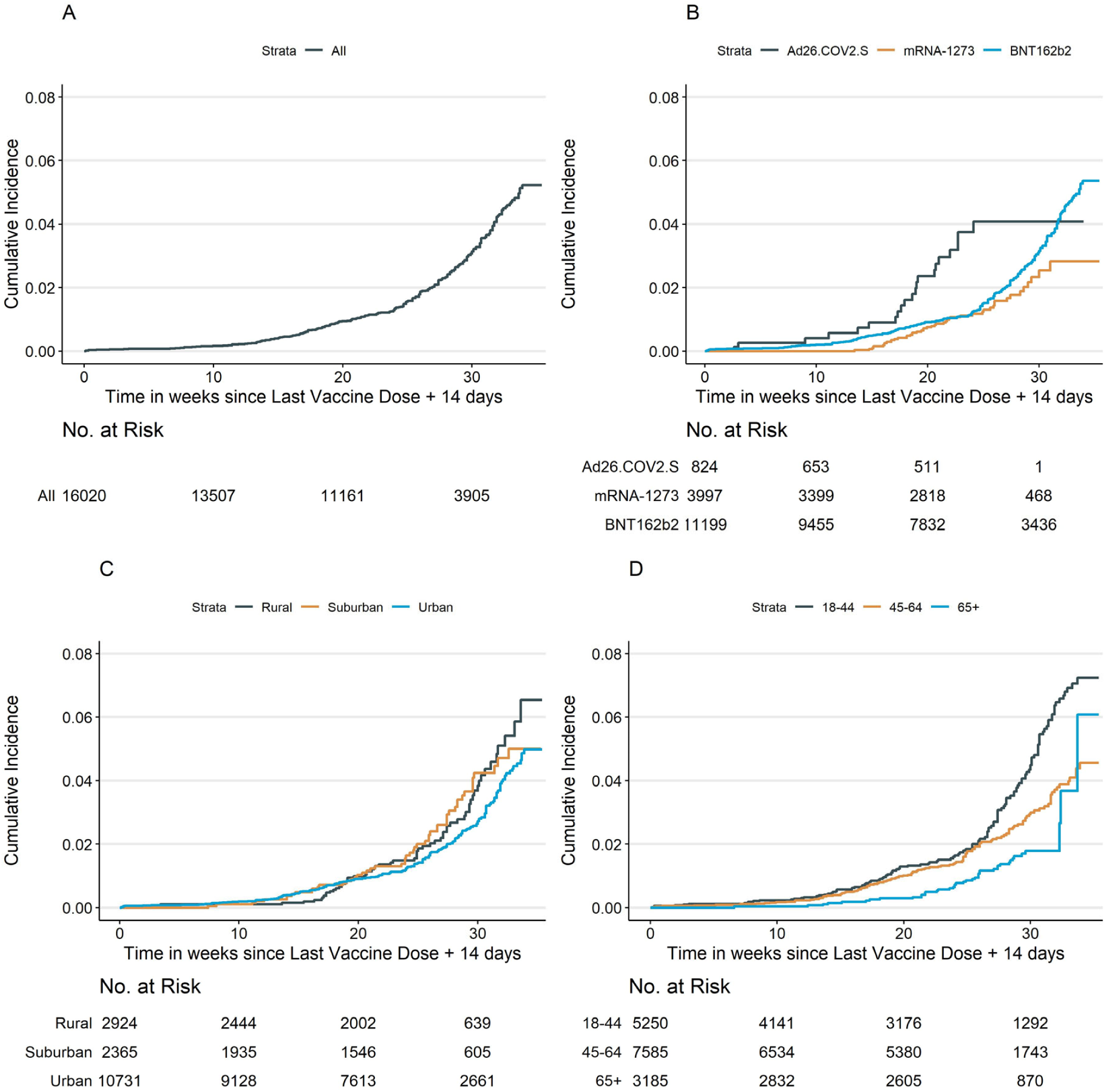
Cumulative Incidence of Self-reported Symptomatic SARS-CoV-2 Test. Cumulative incidence curves (1 minus the unadjusted Kaplan–Meier risk), number at risk and the cumulative number of events at each time point for the first self-reported symptomatic positive SARS-CoV-2 test, starting from 2 weeks after the last dose among participants who reported full vaccination.

## Discussion

In this community-based observational study, the cumulative incidence of participants in North Carolina who self-reported SARS-CoV-2 infection was 5.2% by 34 weeks following full vaccination. The study period included a statewide surge in cases driven by the Delta variant, with a comparable number of new cases as during the winter of 2020-21. The overall breakthrough infection event rate was 7.3 events per 100,000 person-years. Participants vaccinated with Ad26.COV2.S, participants younger than 45 years, and those from rural and suburban counties were at higher risk of breakthrough infection. The association with younger age may reflect more frequent exposures, higher risk exposures, age-associated differences in mitigation behaviors, or a combination of these factors. The association with rural residence may reflect increased transmission in areas with generally lower rates of vaccination ^13-15^ though we did not find a significant difference in breakthrough infection rates when comparing counties with a high (≥ 60%) vs low (< 60%) overall vaccination rate. Race/ethnicity, sex, and occupation as healthcare workers were not associated with higher risk for infection after vaccination. The analysis did not detect a difference in breakthrough infection rates based on general mask usage as a proxy for COVID-19 risk mitigation behavior. However, the conclusions from general mask usage is limited because it is not linked to a specific exposure.

These findings may be limited by selection and reporting biases and may not be generalizable to other areas of the country. Participants were largely recruited through their affiliated healthcare system or EHR (e.g. through electronic patient portal systems); therefore, they may be more engaged in healthcare than the general population. This study includes self-report of a new positive test for COVID-19, generally accompanied by symptoms, so paucisymptomatic or asymptomatic infections may be missed. The results are dependent on transmission rates within the community, which will vary during the course of the pandemic.

In conclusion, this large NC based observational study conducted in a real-world setting demonstrated an event rate of 7.3 breakthrough infections per 100,000 person-years. Breakthrough infection occurred at a higher rate in rural parts of North Carolina, among the younger population and those vaccinated with Ad26.COV2.S. While the underlying causes of the observed differences in breakthrough infection rates deserve further study, our findings suggest that younger adults, those who received the Ad26.COV2.S vaccination, and those residing in suburban and rural counties should be targeted for additional risk mitigation strategies to prevent infection and further transmission of SARS-CoV-2.

## Data Availability

All data produced in the present study are available upon reasonable request to the authors.

## Acknowledgements

The COVID-19 Community Research Partnership gratefully acknowledges the commitment and dedication of the study participants. Programmatic, laboratory, and technical support was provided by Vysnova Partners, Inc., Javara, Inc., and Oracle Corporation.

## Author Contributions

Dr. Uschner and Mr. Bott had full access to all of the data in the study and take responsibility for the integrity of the data and the accuracy of the data analysis.

### Concept and design

Uschner, Santacatterina Herrington, Runyon *Acquisition, analysis, or interpretation of data:* All authors *Drafting of the manuscript:* Uschner, Runyon

### Critical revision of the manuscript for important intellectual content

All authors

### Statistical analysis

Uschner, Bott, Munawar, Seals

### Obtained funding

Herrington, Sanders, Uschner

### Administrative, technical, or material support

Uschner, Bott, Fette, Gunaratne, Burke

### Supervision

Uschner, Santacatterina, Herrington, Sanders, Runyon

## **The COVID-19 Community Research Partnership

### Wake Forest School of Medicine

John Walton Sanders*, MD, MPH, Thomas F Wierzba*, PhD, MPH, David Herrington*, MD, MHS, Mark A. Espeland, PhD, Morgana Mongraw-Chaffin, PhD, Alain Bertoni, MD, Martha A. Alexander-Miller, PhD, Allison Mathews, PhD, Iqra Munawar, MS, Austin Lyles Seals, MS, Brian Ostasiewski, Christine Ann Pittman Ballard, MPH, Metin Gurcan, PhD, Alexander Ivanov, MD, Allison Matthews, PhD, Giselle Melendez Zapata, MD, Marlena Westcott, PhD, Karen Blinson, Laura Blinson, Douglas McGlasson, Mark Mistysyn, Donna Davis, Lynda Doomy, Perrin Henderson, MS, Alicia Jessup, Kimberly Lane, Beverly Levine, PhD, Jessica McCanless, MS, Sharon McDaniel, Kathryn Melius, MS, Christine O’Neill, Angelina Pack, RN, Ritu Rathee, RN, Scott Rushing, Jennifer Sheets, Sandra Soots, RN, Michele Wall, Samantha Wheeler, John White, Lisa Wilkerson, Rebekah Wilson, Kenneth Wilson, Deb Burcombe, John Williamson, PharmD

### Atrium Health

Michael S. Runyon*, MD MPH, Lewis H. McCurdy*, MD, Michael A. Gibbs, MD, Yhenneko J. Taylor, PhD, Lydia Calamari, MD, Hazel Tapp, PhD, Amina Ahmed, MD, Michael Brennan, DDS, Lindsay Munn, PhD RN, Keerti L. Dantuluri, MD, Timothy Hetherington, MS, Lauren C. Lu, Connell Dunn, Melanie Hogg, MS, CCRA, Andrea Price, Marina Leonidas, Laura Staton, Kenniesha Spencer, MPH, Melinda Manning, Whitney Rossman, MS, Frank X. Gohs, MS, Anna Harris, MPH, Bella Gutnik MS, Jennifer S. Priem, PhD, MA, Ryan Burns, MS

### MedStar Health Research Institute

William Weintraub*, MD, Kristen Miller, DrPH, CPPS,, Chris Washington, Allison Moses, Sarahfaye Dolman, Julissa Zelaya-Portillo, John Erkus, Joseph Blumenthal, Ronald E. Romero Barrientos, Sonita Bennett, Shrenik Shah, Shrey Mathur, Christian Boxley, Paul Kolm, PhD, Eva Hochberger, Ella Franklin, Deliya Wesley, Naheed Ahmed

### Tulane

Richard Oberhelman*, MD, Joseph Keating*, PhD, Patricia Kissinger, PhD, John Schieffelin, MD, Joshua Yukich, PhD, Andrew “AJ” Beron, MPH, Devin Hayes, BS, Johanna Teigen, MPH

### University of Maryland School of Medicine

Karen Kotloff*, MD, Wilbur H. Chen*, MD, MS, DeAnna Friedman-Klabanoff, MD, Andrea A. Berry, MD, Helen Powell, PhD, Lynnee Roane, MS, RN, Reva Datar, MPH

### University of Mississippi

Adolfo Correa*, MD, PhD, Leandro Mena*, MD, MPH, Bhagyashri Navalkele, MD, Yuan-I Min, PhD, Alexandra Castillo, MPH, Lori Ward, PhD, MS, Robert P. Santos, MD, Courtney Gomillia, MS-PHS, Pramod Anugu, Yan Gao, MPH, Jason Green, Ramona Sandlin, RHIA, Donald Moore, MS, Lemichal Drake, Dorothy Horton, RN, Kendra L. Johnson, MPH

### Wake Med Health and Hospitals

William H. Lagarde*, MD, LaMonica Daniel, BSCR

### New Hanover

Patrick D. Maguire*, MD, Charin L. Hanlon, MD, Lynette McFayden, RN, Isaura Rigo, MD, Kelli Hines, Lindsay Smith, Alexa Drilling, Monique Harris, Belinda Lissor, Vivian Cook, Maddy Eversole, Terry Herrin, Dennis Murphy, Lauren Kinney, Polly Diehl, Nicholas Abromitis, Tina St. Pierre, Judy Kennedy BSCS, MBA, Lauren Kinney, BS, Bill Heckman, Denise Evans, Vivian Cook, Maddy Eversole, Julian March, Ben Whitlock, Wendy Moore

### Vidant Health

Thomas R. Gallaher*, MD, Shakira Henderson, PhD, DNP, MS, MPH, Michael Zimmer, PhD, Danielle Oliver, Tina Dixon, Kasheta Jackson, Martha Reavis, Monica Menon, Brandon Bishop, Rachel Roeth, Mathew Johanson, Alesia Ceaser, Amada Fernandez, Carmen Williams, Jeremiah Hargett, Keeaira Boyd, Kevonna Forbes, Latasha Thomas, Markee Jenkins, Monica Coward, Derrick Clark, Omeshia Frost, Angela Darden, Lakeya Askew, Sarah Phipps, Victoria Barnes

### Campbell University School of Osteopathic Medicine

Robin King-Thiele*, DO, Terri S. Hamrick*, PhD, Chika Okafor, MD (Cape Fear Valley Medical Center), Regina B. Bray Brown, MD (Harnett Health System, Inc.), Pinoorma Vinod, MD (Southeastern Health), Lawrence Klima, MD (Harnett Health System), Amber Brewster, MD (Harnett Health System), Danius Bouyi, DO (Harnett Health System), Katrina Lamont, MD (Harnett Health System), Kazumi Yoshinaga, DO (Harnett Health System), Abdalla Ihmeidan, MHA (Campbell University School of Osteopathic Medicine), A. Suman Peela, MD (Southeastern Health System), Giera Denbel, MD (Southeastern Health System), Jason Lo, MD: Southeastern Health System, Mariam Mayet-Khan, DO (Southeastern Health System), Akash Mittal, DO (Southeastern Health System), Reena Motwani, MD (Southeastern Health System), Mohamed Raafat, MD (Southeastern Health System), Evan Schultz, DO (Cumberland County Hospital System, Cape Fear Valley), Aderson Joseph, MD (Cumberland County Hospital System, Cape Fear Valley), Aalok Parkeh, DO (Cumberland County Hospital System, Cape Fear Valley), Dhara Patel, MD (Cumberland County Hospital System, Cape Fear Valley), Babar Afridi, DO (Cumberland County Hospital System, Cape Fear Valley)

### George Washington University Data Coordinating Center

Diane Uschner*, PhD, Sharon L Edelstein, ScM, Michele Santacatterina, PhD, Greg Strylewicz, PhD, Brian Burke, MS, Mihili Gunaratne, MPH, Meghan Turney, MA, Shirley Qin Zhou, MS, Ashley H Tjaden, MPH, Lida Fette, MS, Asare Buahin, Matthew Bott, Sophia Graziani, Ashvi Soni, MS

### George Washington University Mores Lab

Christopher Mores, PhD, Abigail Porzucek, MS

### Oracle Corporation

Rebecca Laborde

### Vysnova Partners

Anne McKeague, PhD, Grace Tran, MPH, Johnathan Ward, Joyce Dieterly, MPH, Nana Darko, MPH, Kimberly Castellon, Isabella Malcolm, Ryan Brink, MS, Haris Shehzad, MS, Derek Kuprianov

### Sneez LLC

Lucy Guill, MBA, Danielle Lamphier, MBA, Anna Schaefer, MSM, William M. Satterwhite, JD, MD

### Javara Inc

Atira Goodwin

### External Advisory Council

Ruth Berkelman, MD, Emory, Kimberly Hanson, MD, U of Utah, Scott Zeger, PhD, Johns Hopkins, Cavan Reilly, PhD, U. of Minnesota, Kathy Edwards, MD, Vanderbilt, Helene Gayle, MD MPH, Chicago Community Trust

(*Site Principal Investigator)

## Online-Only Figures

**eFigure 1.**
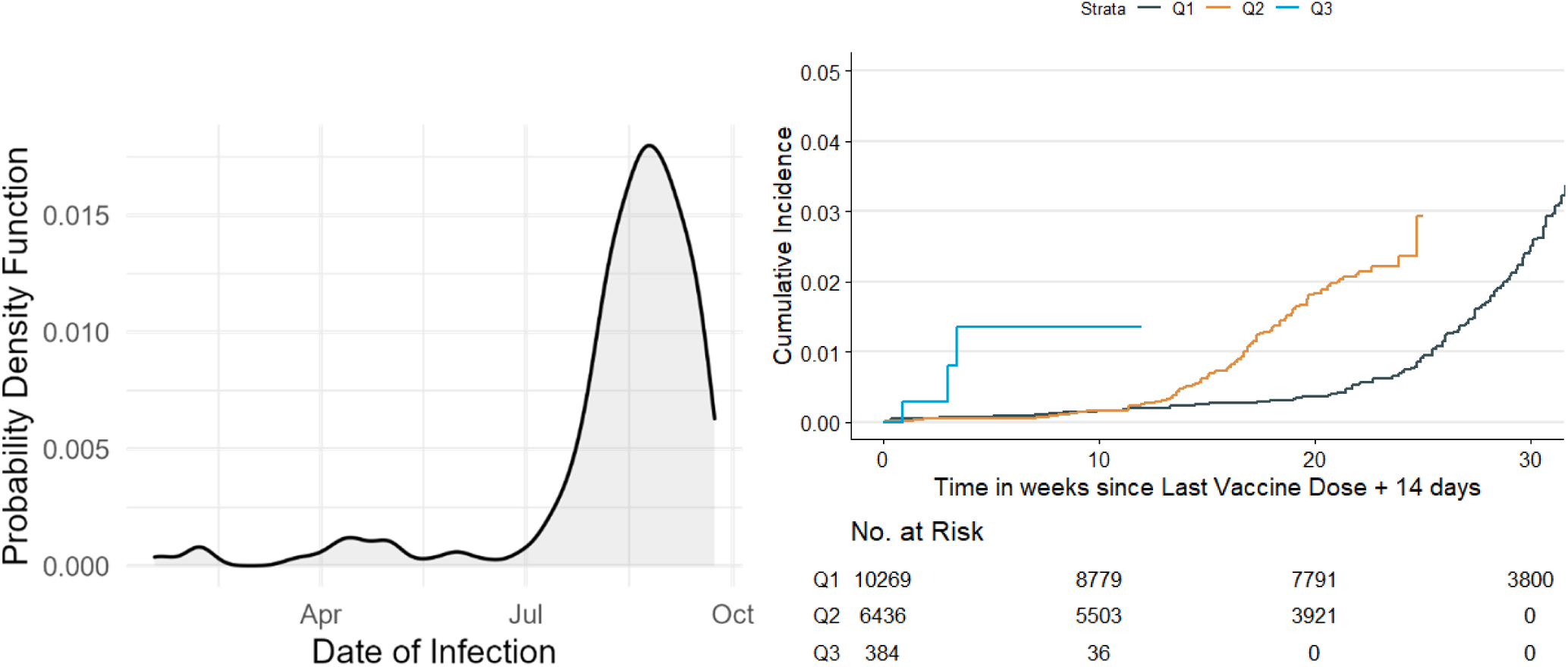
SARS-CoV-2 Infections after vaccination and calendar time. Distribution of SARS-CoV-2 infections after vaccinations by time (LHS) and cumulative incidence by quarter of vaccination date (RHS, Q1 = Vaccination in January-March 2021, Q2 = Vaccination in April-June 2021, Q3 = Vaccination in July-September 2021)

**eFigure 2.**
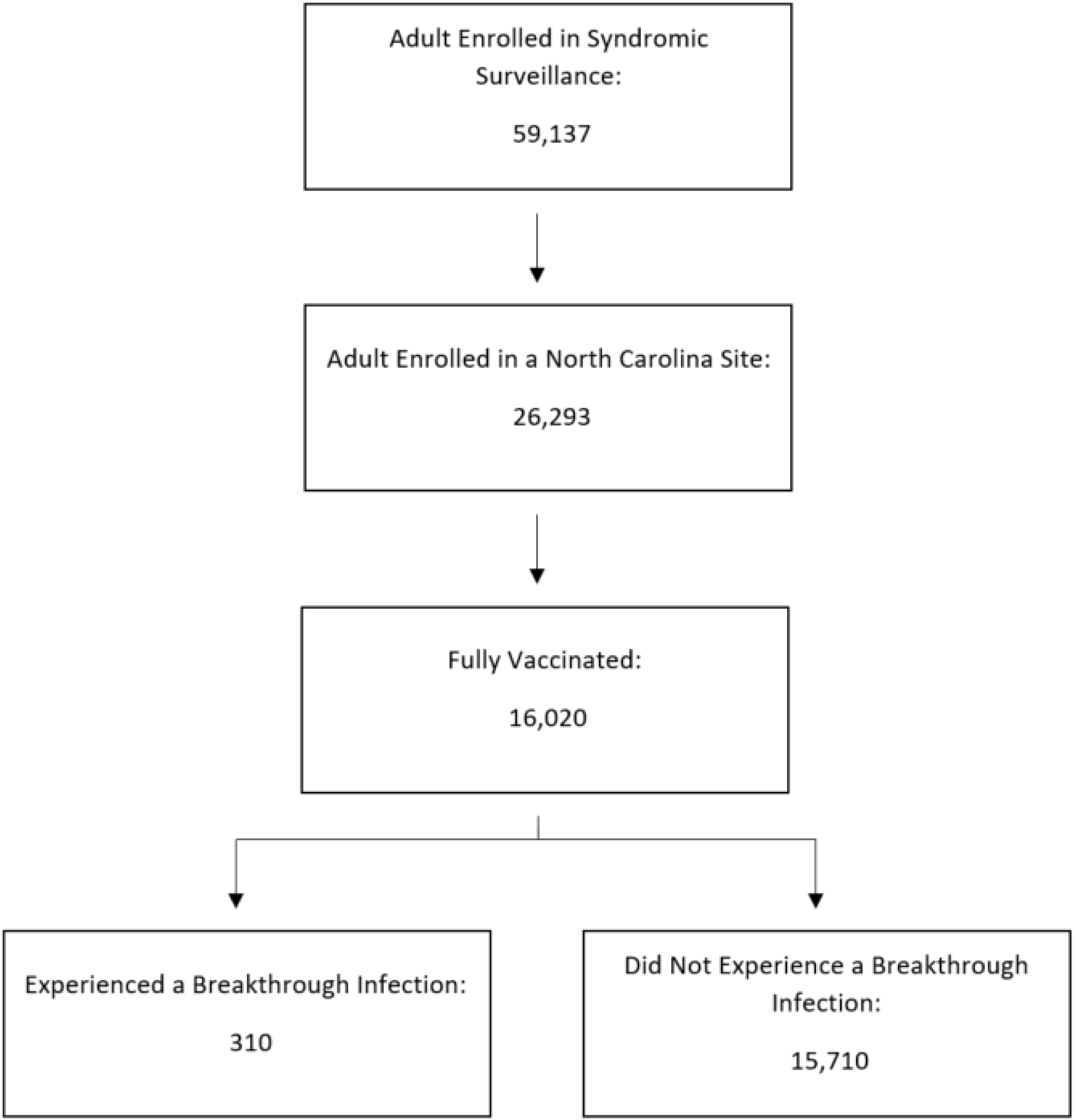
Flow diagram describing the study population. NA

